# Development, validation, and usage of metrics to evaluate the quality of clinical research hypotheses

**DOI:** 10.1101/2023.01.17.23284666

**Authors:** Xia Jing, Yuchun Zhou, James J. Cimino, Jay H. Shubrook, Vimla L. Patel, Sonsoles De Lacalle, Aneesa Weaver, Chang Liu

## Abstract

**Objectives:** Metrics and instruments can provide guidance for clinical researchers to assess their potential research projects at an early stage before significant investment. Furthermore, metrics can also provide structured criteria for peer reviewers to assess others’ clinical research manuscripts or grant proposals. This study aimed to develop, test, validate, and use evaluation metrics and instruments to accurately, consistently, and conveniently assess the quality of scientific hypotheses for clinical research projects.

**Materials and Methods:** Metrics development went through iterative stages, including literature review, metrics and instrument development, internal and external testing and validation, and continuous revisions in each stage based on feedback. Furthermore, two experiments were conducted to determine brief and comprehensive versions of the instrument.

**Results:** The brief version of the instrument contained three dimensions: validity, significance, and feasibility. The comprehensive version of metrics included novelty, clinical relevance, potential benefits and risks, ethicality, testability, clarity, interestingness, and the three dimensions of the brief version. Each evaluation dimension included 2 to 5 subitems to evaluate the specific aspects of each dimension. For example, validity included clinical validity and scientific validity. The brief and comprehensive versions of the instruments included 12 and 39 subitems, respectively. Each subitem used a 5-point Likert scale.

**Conclusion:** The validated brief and comprehensive versions of metrics can provide standardized, consistent, and generic measurements for clinical research hypotheses, allow clinical researchers to prioritize their research ideas systematically, objectively, and consistently, and can be used as a tool for quality assessment during the peer review process.

## Introduction

A hypothesis is an educated guess or statement about the relationship between two or more variables [1,2]. The hypothesis generation process is critical and decisive in determining the significance of a clinical research project or scientific project. Although much progress has been achieved in scientific thinking, reasoning, and analogy [3-8], which are critical skills in hypothesis generation, knowledge about the scientific hypothesis generation process, including how to facilitate the process, especially in a clinical research context, is limited. Many data science researchers believe that secondary data analytic tools can facilitate hypothesis generation [9]. Nevertheless, there is a lack of studies demonstrating the role of a secondary data analysis tool in this process in clinical research. We developed a visual interactive analytic tool for filtering and summarizing large health data sets coded with hierarchical terminologies (VIADS, https://www.viads.info [10]) to filter, compare, summarize, and visualize datasets coded with hierarchical terminologies (e.g., International Classification of Diseases, 9^th^ Revision, Clinical Modification, ICD-9-CM). VIADS can also assist clinical researchers with generating hypotheses. Visual examples of VIADS include hierarchical graphs, bar charts, and 3D plots. Users can obtain expanded information via interactive features, change graph layouts (e.g., small, medium, and large horizontal spacing), zoom in and out, and move, save, and export graphs and their data files.

To put this manuscript in the appropriate context, we provide some background information on the entire project and how we conducted it to elaborate on how it fits the bigger picture. To explore the clinical researchers’ hypothesis generation processes, we conducted one-on-one study sessions in which researchers (i.e., participants) generated hypotheses using the same datasets within two hours with or without VIADS [11]. This was a 2 × 2 study design (with and without VIADS by experienced and inexperienced clinical researchers). The quality of each scientific hypothesis generated by the participants in the study [12,13] was assessed by an expert panel using the same metrics. The aggregated quality assessment results, along with the number of hypotheses and the average time used to generate a hypothesis, were used to detect the differences in the hypotheses generated by the participants [12]. A reliable, generic, and convenient tool is required to have a reliable, consistent, and accurate assessment of the quality of the generated scientific hypotheses [14].

The original purpose of developing metrics is to evaluate the hypotheses generated by the participants in our research project. Furthermore, the validated metrics and instruments can be very useful in clinical research. Researchers can use the instruments to compare and select more valuable and impactful hypotheses to pursue in their research endeavor at an early stage before any significant investment in resources. Furthermore, the instruments can be used during peer review processes for clinical research manuscripts or grant proposals. Traditionally, the peer review process is conducted by human experts, which can be a subjective assessment. Using an explicit, clearly defined, consistent, and comprehensive assessment tool based on metrics can provide a solid foundation for a relatively more objective, consistent, and perhaps more accurate evaluation during the peer review process of clinical research projects. The lack of a significant, meaningful, and impactful hypothesis to start with can make all other aspects of the research projects meaningless, regardless of rigor or validity. Therefore, the development and validation of such metrics play an important role in facilitating the launch of a more impactful research project and conducting a more objective, consistent, and accurate peer-review evaluation. In this manuscript, we introduce the approach we used to develop and validate the metrics, the results of the metrics and instruments, and the preliminary experience of the usage of the metrics. We hope to share the metrics and instruments as potential tools and the methodology we used to develop them with the clinical research community.

## Materials and Methods

### Study flow and internal validation

The development of the metrics went through a series of iterative stages (Figure 1) [15-17]. One author (XJ, a medical informatics researcher) reviewed the literature and drafted the metrics. Then, two authors (XJ and YCZ, a research methodologist) discussed the outlined metrics, formulated the initial metrics, and revised the metrics after all confusion and concerns were addressed. This was the first internal validation level between two team members. The adjusted metrics were distributed to the research team as anonymous surveys about the evaluation items for feedback. This step was conducted in three rounds to incorporate all the feedback received. This step constituted the second level of internal validation among the entire team.

**Figure 1.**
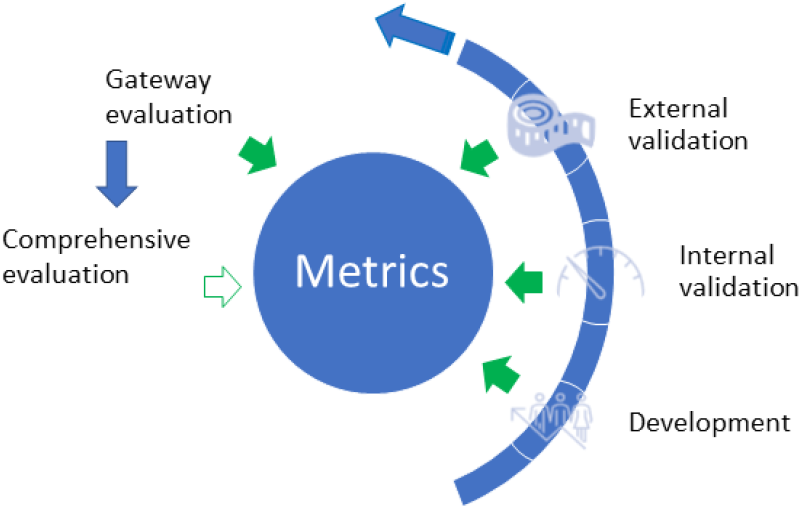
Development, validation, and usage of the metrics to assess the quality of clinical research hypotheses. Blue arrows indicate the development stages of metrics; solid green arrows indicate the feedback incorporated into the metrics from each stage; green hollowed arrow indicates future work

### External validation

After completing the internal validation, an iterative external validation process was conducted by engaging additional four invited clinical research experts. The criteria to be eligible as a clinical research expert were pre-defined during the design of the research project (please refer to our prior publication for details [11]). The instrument used in the initial external validation is shown in Appendix 1. The internal validation processes on the instrument (i.e., the evaluation dimensions, subitems, and scales of subitems) followed a revised Delphi method [18-22], which included transparent and open discussions (via face-to-face meetings, emails, and complementary video conferences) among the research team.

The external validation consisted of three steps, (1) initial validation of the metrics, (2) experimental evaluations by using the metrics to assess hypotheses generated during the study sessions, and (3) refinement based on the feedback and results of the experimental evaluations (Figure 2). A survey (Appendix 2) that served as the medium validation instrument was used among all expert panel members (including three senior consultants from the research team and four external clinical research experts) to obtain feedback, which was incorporated into the final metrics (Table 1 and Appendix 3). A 10-item evaluation instrument was formulated from the development and validation processes.

**Figure 2.**
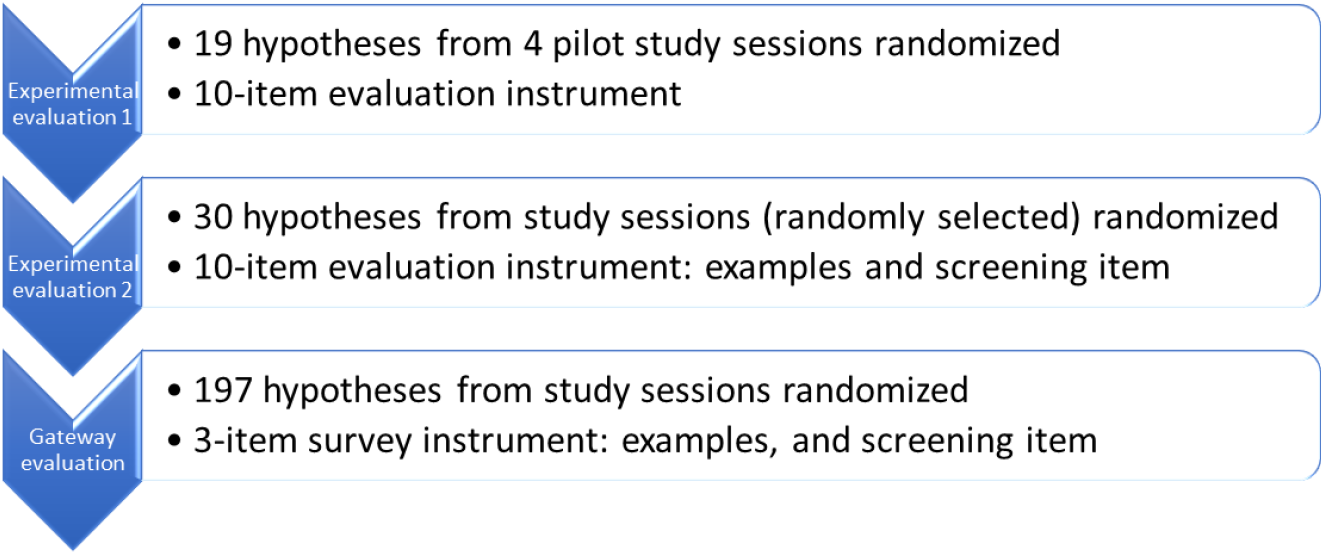
Refinement process of the clinical research hypotheses quality evaluation instrument

**Table 1.**
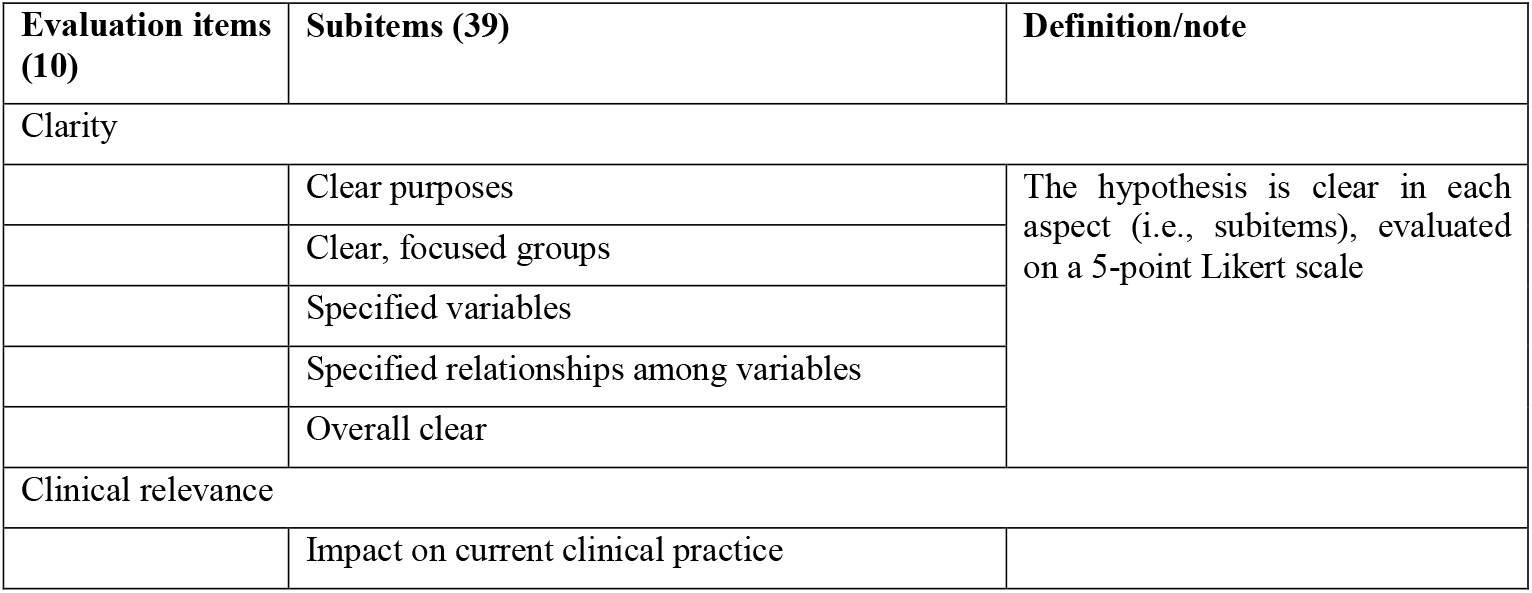
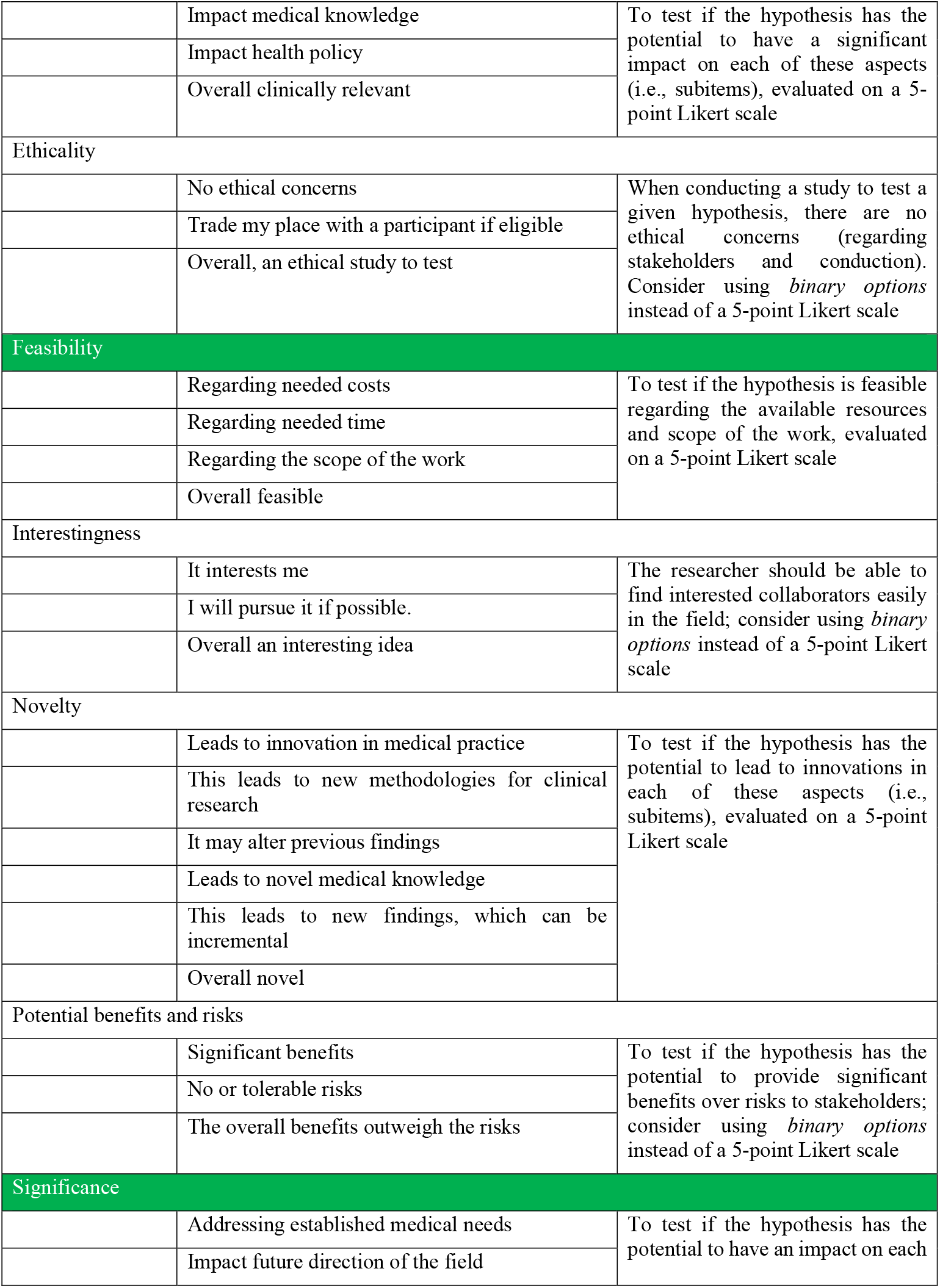
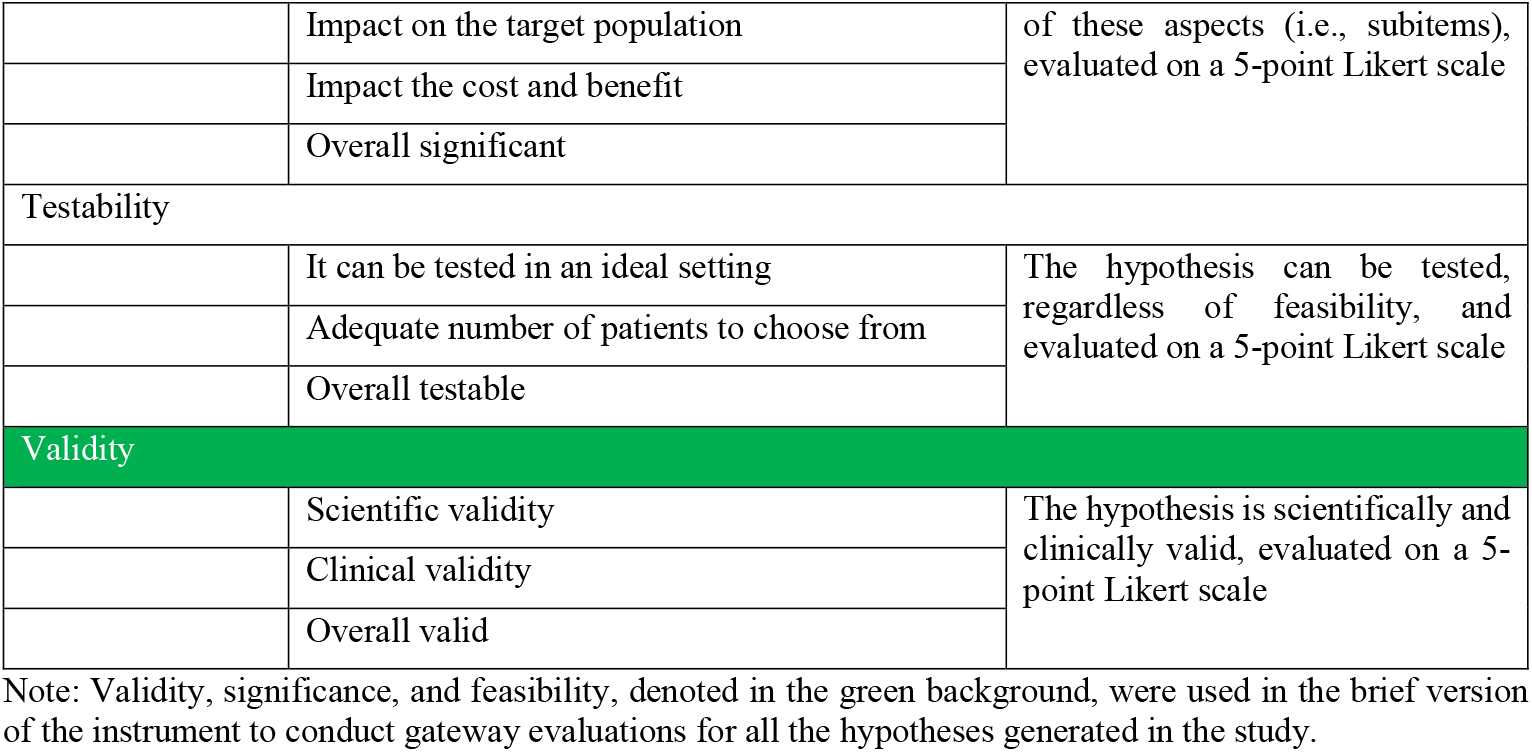
Evaluation items and subitems in the metrics used to assess the scientific hypotheses in clinical research

| Evaluation items (10) | Subitems (39) | Definition/note |
| --- | --- | --- |
| Clarity |  |  |
|  | Clear purposes | The hypothesis is clear in each aspect (i.e., subitems), evaluated on a 5-point Likert scale |
|  | Clear, focused groups |  |
|  | Specified variables |  |
|  | Specified relationships among variables |  |
|  | Overall clear |  |
| Clinical relevance |  |  |
|  | Impact on current clinical practice |  |
|  | Impact medical knowledge | To test if the hypothesis has the potential to have a significant impact on each of these aspects (i.e., subitems), evaluated on a 5-point Likert scale |
|  | Impact health policy |  |
|  | Overall clinically relevant |  |
| Ethicality |  |  |
|  | No ethical concerns | When conducting a study to test a given hypothesis, there are no ethical concerns (regarding stakeholders and conduction). Consider using *binary options* instead of a 5-point Likert scale |
|  | Trade my place with a participant if eligible |  |
|  | Overall, an ethical study to test |  |
| Feasibility |  |  |
|  | Regarding needed costs | To test if the hypothesis is feasible regarding the available resources and scope of the work, evaluated on a 5-point Likert scale |
|  | Regarding needed time |  |
|  | Regarding the scope of the work |  |
|  | Overall feasible |  |
| Interestingness |  |  |
|  | It interests me | The researcher should be able to find interested collaborators easily in the field; consider using *binary options* instead of a 5-point Likert scale |
|  | I will pursue it if possible. |  |
|  | Overall an interesting idea |  |
| Novelty |  |  |
|  | Leads to innovation in medical practice | To test if the hypothesis has the potential to lead to innovations in each of these aspects (i.e., subitems), evaluated on a 5-point Likert scale |
|  | This leads to new methodologies for clinical research |  |
|  | It may alter previous findings |  |
|  | Leads to novel medical knowledge |  |
|  | This leads to new findings, which can be incremental |  |
|  | Overall novel |  |
| Potential benefits and risks |  |  |
|  | Significant benefits | To test if the hypothesis has the potential to provide significant benefits over risks to stakeholders; consider using *binary options* instead of a 5-point Likert scale |
|  | No or tolerable risks |  |
|  | The overall benefits outweigh the risks |  |
| Significance |  |  |
|  | Addressing established medical needs | To test if the hypothesis has the potential to have an impact on each |
|  | Impact future direction of the field |  |
|  | Impact on the target population | of these aspects (i.e., subitems),<br>evaluated on a 5-point Likert scale |
|  | Impact the cost and benefit |  |
|  | Overall significant |  |
| Testability |  |  |
|  | It can be tested in an ideal setting | The hypothesis can be tested,<br>regardless of feasibility, and<br>evaluated on a 5-point Likert scale |
|  | Adequate number of patients to choose from |  |
|  | Overall testable |  |
| Validity |  |  |
|  | Scientific validity | The hypothesis is scientifically and<br>clinically valid, evaluated on a 5-point Likert scale |
|  | Clinical validity |  |
|  | Overall valid |  |

Note: Validity, significance, and feasibility, denoted in the green background, were used in the brief version of the instrument to conduct gateway evaluations for all the hypotheses generated in the study.

### Experimental evaluation 1

In experimental evaluation 1, we performed validation analysis for the ten evaluation items (without subitems) using 19 hypotheses generated via pilot studies of the research project. These hypotheses were randomly assigned to two Qualtrics surveys (10 and 9 hypotheses). The seven members are our evaluation team (i.e., an expert panel), all of whom with a medical or methodology background. They rated all the hypotheses. The inter-rater agreement of the seven experts’ ratings on the 19 hypotheses was analyzed using the intra-class correlation (ICC). We used descriptive statistics to analyze the results of the survey. Based on the mean results (i.e., the average rating scores for each hypothesis) from experimental evaluation 1, we identified the best and worst examples of hypotheses, which were used in experimental evaluation 2 to provide examples for the expert panel members to better calibrate their rating scores in the assessment of the remaining hypotheses.

### Experimental evaluation 2

Experimental evaluation 2 included 30 randomly selected hypotheses from the study sessions using the 10-item evaluation instrument (Figure 3). In the instructions, we provided the best and worst examples of hypotheses based on the experimental evaluation 1 results and set a screening item: validity. If a statement is not a hypothesis, further evaluation is unnecessary. If three or more experts scored at 1 (lowest rating) in validity for any of the hypotheses, it was removed from the following analysis. ICC analysis was performed to examine the consistency of the seven experts’ ratings on the valid hypotheses using the ten items. The evaluation results were compared using a paired *t*-test analysis.

**Figure 3.**
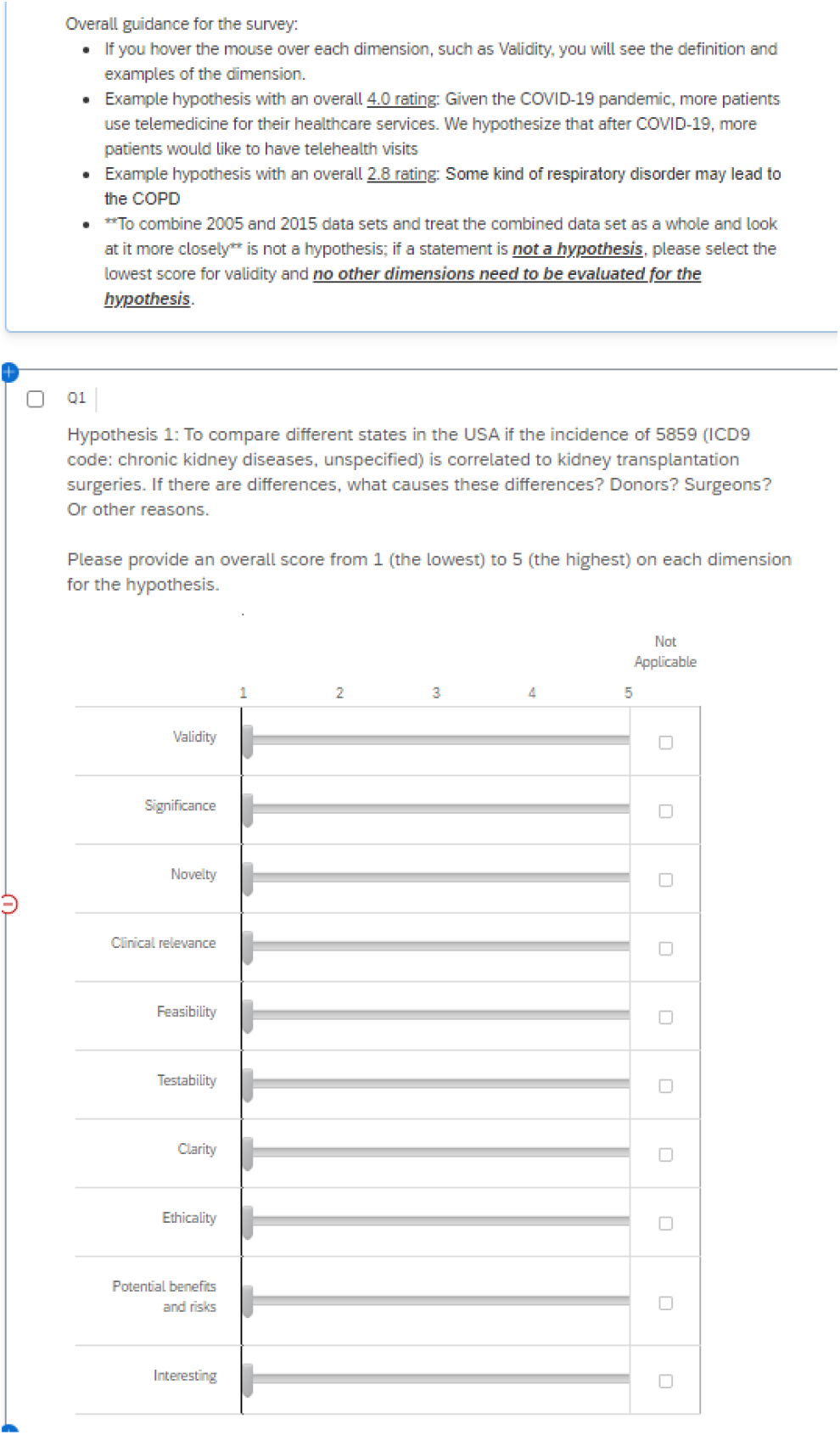
Ten-item evaluation instrument for clinical research hypothesis screening and evaluation

### Instruments used for validation and experiment evaluations

All steps mentioned above (initial draft development, internal validation, external validation, refinement, and revisions in between the steps) were conducted iteratively using quantitative and qualitative approaches (e.g., Qualtrics surveys, emails, additional phone calls, and virtual conferences). The evaluations of the instrument (with 10 items and 39 subitems), i.e., the validation process before experts used the instrument to conduct the experimental evaluations, including a 5-point Likert scale and three additional options of unable to assess, unnecessary subitem, or use this item only (Appendix 2). The evaluation instrument (with 10 items) used in experimental evaluations 1 and 2 included a 5-point Likert scale and an option of not applicable (Figure 3). The gateway evaluation and the results are published separately. This study was approved by the Ohio University Institutional Review Board (18-X-192) and Clemson University Institutional Review Board (IRB2020-056).

## Results

We present comprehensive (10 items and 39 subitems, Appendix 3) and brief versions (3 items, 12 subitems, Table 1, Appendix 4) of the instrument to assess the quality of clinical research hypotheses and the evidence generated from experimental evaluations. Most measurements for evaluating the quality of clinical research hypotheses from the literature [1,2,9,23-33] include the following ten dimensions: ***validity, significance, novelty, clinical relevance, potential benefits and risks, ethicality, feasibility, testability, clarity***, and ***researcher interest level***. We developed 39 sub-items to measure each dimension comprehensively and unambiguously (Table 1). The quality of each item was measured using a 5-point Likert scale. Table 1 shows all the evaluation items and subitems and how they were used to evaluate the quality of clinical research hypotheses. Table 2 presents two examples of hypotheses and their quality evaluation results among all evaluators when using the 3-item instrument (Appendix 4).

**Table 2.**
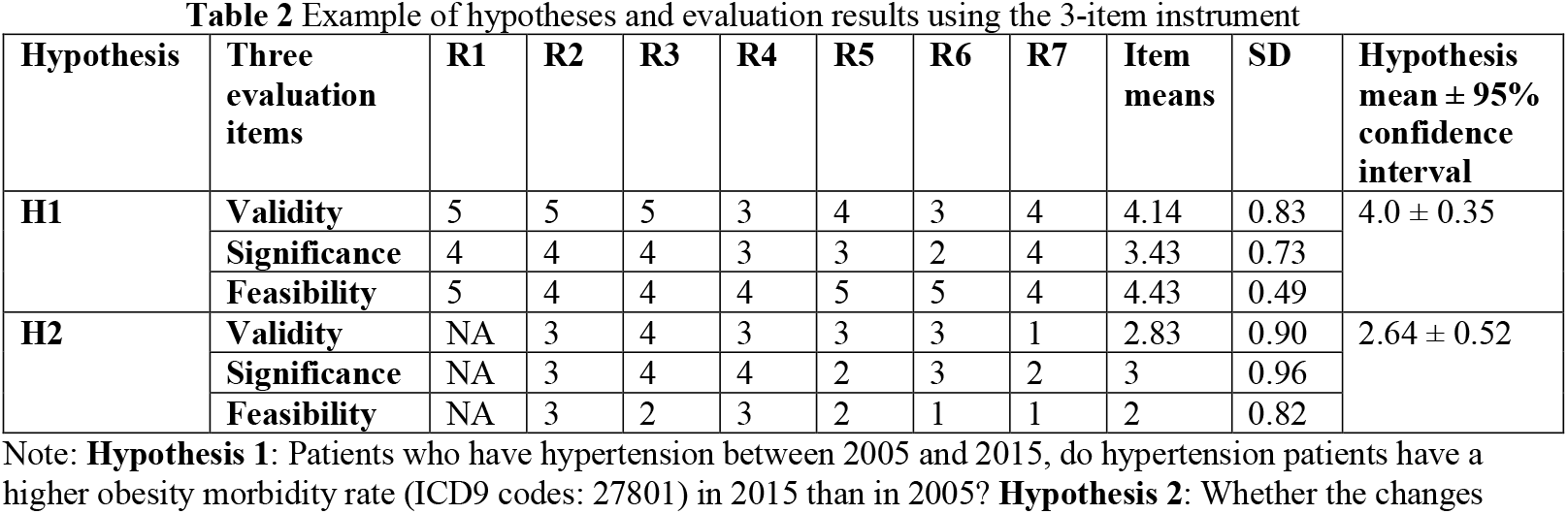
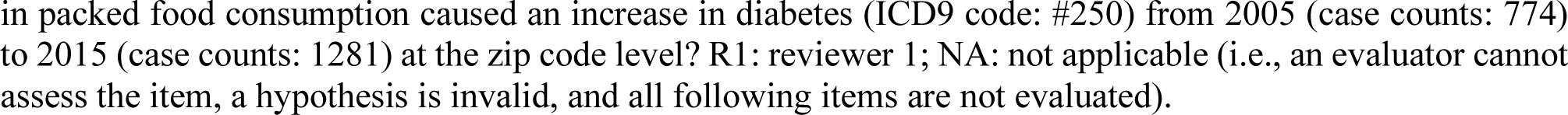
Example of hypotheses and evaluation results using the 3-item instrument

| Hypothesis | Three evaluation items | R1 | R2 | R3 | R4 | R5 | R6 | R7 | Item means | SD | Hypothesis mean ± 95% confidence interval |
| --- | --- | --- | --- | --- | --- | --- | --- | --- | --- | --- | --- |
| **H1** | **Validity** | 5 | 5 | 5 | 3 | 4 | 3 | 4 | 4.14 | 0.83 | 4.0 ± 0.35 |
|  | **Significance** | 4 | 4 | 4 | 3 | 3 | 2 | 4 | 3.43 | 0.73 |  |
|  | **Feasibility** | 5 | 4 | 4 | 4 | 5 | 5 | 4 | 4.43 | 0.49 |  |
| **H2** | **Validity** | NA | 3 | 4 | 3 | 3 | 3 | 1 | 2.83 | 0.90 | 2.64 ± 0.52 |
|  | **Significance** | NA | 3 | 4 | 4 | 2 | 3 | 2 | 3 | 0.96 |  |
|  | **Feasibility** | NA | 3 | 2 | 3 | 2 | 1 | 1 | 2 | 0.82 |  |

Note: **Hypothesis 1:** Patients who have hypertension between 2005 and 2015, do hypertension patients have a higher obesity morbidity rate (ICD9 codes: 27801) in 2015 than in 2005? **Hypothesis 2:** Whether the changes
in packed food consumption caused an increase in diabetes (ICD9 code: #250) from 2005 (case counts: 774) to 2015 (case counts: 1281) at the zip code level? R1: reviewer 1; NA: not applicable (i.e., an evaluator cannot assess the item, a hypothesis is invalid, and all following items are not evaluated).

In experimental evaluation 1, the experts’ evaluation scores for the 19 hypotheses across the ten criteria were averaged, and none of the ten criteria could achieve a moderate ICC coefficient (>0.50). Therefore, experimental evaluation 2 was conducted, validity was set as a screening item, and one best and one not-so-good examples of hypotheses from experimental evaluation 1 were provided in the instructions of experimental evaluation 2.

In the experimental evaluation 2 result analysis, the results of the screening item were checked first. The valid sample size included 17 hypotheses (out of 30) in experimental evaluation 2. Then, the inter-rater agreement of the 17 hypotheses was checked using ICC analyses. Half the ten criteria achieved a moderate ICC value (0.50–0.75). Based on the ICC results and qualitative evaluation of the ten criteria, a decision was made to retain three measures (i.e., validity, significance, and feasibility) for a shortened version of the evaluation instrument.

The paired *t*-test indicated no significant difference (t = 1.74, p = .13) between the ratings using the 3-item (Appendix 4) and 10-item instruments. Figure 3 shows the 10-item evaluation instrument used for experimental evaluation 2, including the best and worst examples. Figure 4 presents the steps used in this study and the corresponding results to provide a summary view of the methods and results.

**Figure 4.**
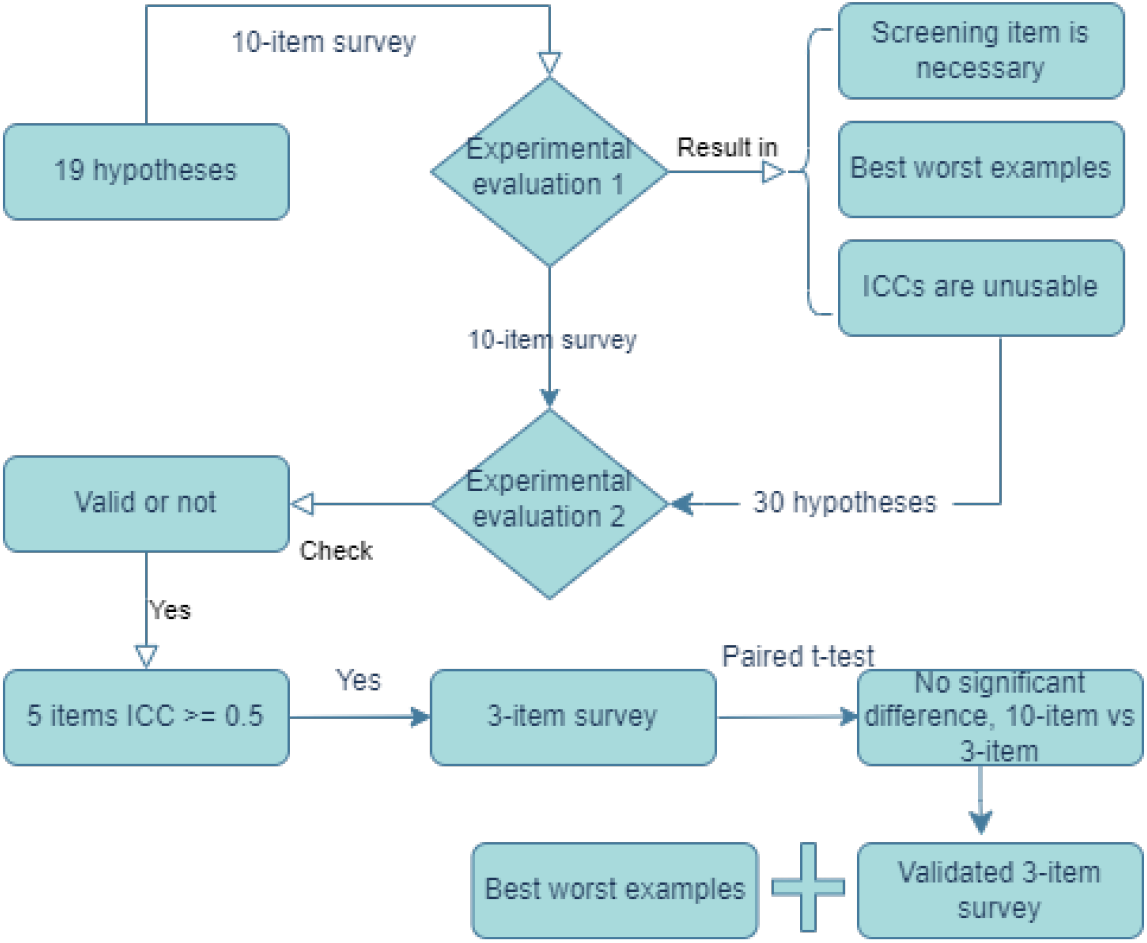
Summary of methods, steps, and corresponding results of development and validation of metrics in assessing the quality of clinical research hypotheses

## Discussion

Hypothesis generation is a highly sophisticated cognitive process. Not all information used during the processes is a conscious or explicit choice. Our study explored the process of scientific hypothesis generation using the same clinical datasets to determine whether a secondary data analytic tool could facilitate the process. Establishing the evaluation metrics was the first step and was the critical foundation for the overall study and understanding of the entire process. Comprehensive and objective measures were given more weight during the development of the metrics. In our studies, the clinical researchers generated a few to over a dozen hypotheses within two hours [12,13]. However, not all hypotheses were of high quality. Therefore, it was not conducive to using the experts’ time to comprehensively evaluate each hypothesis generated during the study sessions.

Furthermore, using the entire set of metrics, including all items and subitems, to evaluate each generated hypothesis may be unnecessary. Thus, we used “gateway” evaluations as a filter to identify the higher-quality hypotheses. The experts can determine the higher-quality hypotheses more carefully, thoroughly, and comprehensively during the comprehensive evaluation. Therefore, validity was used as a screening item, and the “not a hypothesis” option was added in the initial assessment, enlightened by the experimental evaluation 1 results.

The results of experimental evaluation 2 aided in determining a brief evaluation instrument with the 3 items used to evaluate the rest of the hypotheses generated by the participants during the gateway evaluation (Figures 1 and 2). From the ICC analysis in experimental evaluation 2, feasibility, testability, and clarity have the highest ICC values among the ten items. However, empirically, we highly prioritize validity, significance, and novelty. Combining our experience and the statistical testing results, we developed two options: validity, significance, and feasibility; validity, significance, clinical relevance, and feasibility. The testing results indicated that both were valid options. Thus, we determined the 3-item evaluation instrument for operational purposes. We used our experience and the statistical testing results to make the decision.

Meanwhile, we noticed negative ICC values in ethicality, potential benefits and risks, and interestingness. The results indicated that reaching a consensus on these items might be challenging. We recommend that these three items change to a binary (yes/no) category instead of a 5-point Likert scale to simplify the evaluation and improve the agreement among the evaluators.

During the external validation, one major result was to add “not applicable” as an option to the evaluation instrument under each item and subitem. Considering the different backgrounds of expert panel members, this additional option helped them to simplify the evaluation process. Comparing the statistical results, we noticed a significant improvement in experimental evaluation 2, mainly due to the examples of the best and worst hypotheses, which might assist evaluators in calibrating their expectations. Furthermore, we reminded the evaluators that some statements were not hypotheses, i.e., we used validity as a screening item. The experimental evaluation 2 results are based on 17 valid hypotheses. The 13 invalid hypotheses have three or more expert panel members who evaluated them as 1 (the lowest score) in validity.

Although the evaluation of a particular hypothesis by an expert can be subjective, we used examples of the best and worst hypotheses to assist experts in calibrating their expectations more accurately. The inclusion of seven expert members balances the subjectivity and provides a more consistent evaluation using the same instrument. In addition, we used objective measures, e.g., the number of hypotheses generated and the average time spent on each hypothesis, and randomized the hypotheses during the assessment. These strategies helped the expert panel to provide more consistent evaluations and allowed us to accurately conclude the quality of the hypotheses.

## Conclusion

The metrics and instruments developed in this study can benefit clinical researchers in evaluating their hypotheses more comprehensively, consistently, and efficiently before launching a research project, as well as providing valid instruments for the peer review process in clinical research. Our results provide an evidence-based brief version (validity, significance, and feasibility) and a comprehensive version of the evaluation items (validity, significance, feasibility, novelty, clinical relevance, testability, clarity, ethicality, potential benefits and risks, and interesting to others) to assess the quality of clinical research hypotheses. The metrics can be used to standardize the process and provide a consistent tool for this highly sophisticated cognitive process.

## Supporting information

The instrument used in the initial external validation is shown in Appendix 1.

A survey (Appendix 2) that served as the medium validation instrument was used among all expert panel members

which was incorporated into the final metrics (Table 1 and Appendix 3).

We present comprehensive (10 items and 39 subitems, Appendix 3) and brief versions (3 items, 12 subitems, Table 1, Appendix 4)

## Data Availability

All data produced in the present study are available upon reasonable request to the correspondence author.

---

**Appendix 1**: Initial survey instrument used for external validation of the evaluation items

**Appendix 2**: Medium survey instrument used for external validation of the evaluation items

**Appendix 3**: Evaluation instrument of 10 items with subitems (full version) to evaluate the scientific hypotheses in clinical research

**Appendix 4:** Evaluation instrument of 3 items without subitems to evaluate the scientific hypotheses in clinical research

## Acknowledgments and funding

The project was supported by the National Library of Medicine (R15LM012941) and partially supported by the National Institute of General Medical Sciences of the United States National Institutes of Health (P20GM121342).

