## Supplementary material for "Development, validation, and usage of metrics to evaluate the quality of clinical research hypotheses": The instrument used in the initial external validation is shown in Appendix 1.

### VIADS_Expert_Metrics_Feedback20200616

Start of Block: Default Question Block

This is a feedback survey about the rubrics that you will use to evaluate each hypothesis generated by VIADS experiment participants. Your comments and suggestions on each evaluation item (in blue font) in the rubrics are greatly appreciated!

Example hypothesis:
1) Drug A and drug B are similar in treating disease C. After analyzing data from 2017 in institution X; we notice Drug A was associated with adverse cardiovascular events at a rate significantly higher than with Drug B.

a. Hypothesis: Analysis of date from 2017 in institution Y will show similar higher associations with disease C for Drug A than Drug B.

Q1.1 The **validity** of this hypothesis will be evaluated by the following metrics (the metrics are for text display purpose, no need to answer):

| | Strongly disagree (1) | Disagree (2) | Neutral (3) | Agree (4) | Strongly agree (5) |
|---|---|---|---|---|---|
| The hypothesis is valid **scientifically** (1) | ○ | ○ | ○ | ○ | ○ |
| The hypothesis is valid **clinically**, i.e., sound basis clinically (2) | ○ | ○ | ○ | ○ | ○ |

Q1.2 Can the validity of the hypothesis be evaluated clearly by the above statements?

○ Yes (1)

○ No (2)

---

Q1.3 Are there any additional items that should be included to measure the validity of the hypothesis?

________________________________________________________________

________________________________________________________________

________________________________________________________________

________________________________________________________________

________________________________________________________________

---

Q1.4 Do you have any additional comments about the evaluation of the validity of the hypothesis?

________________________________________________________________

________________________________________________________________

________________________________________________________________

________________________________________________________________

________________________________________________________________

---

Q1.5 The potential benefits, risks of this hypothesis will be evaluated by the following metrics (the metrics are for text display purpose, no need to answer):

| | Strongly disagree (1) | Disagree (2) | Neutral (3) | Agree (4) | Strongly agree (5) |
|---|---|---|---|---|---|
| The hypothesis will bring significant **benefits** (1) | ○ | ○ | ○ | ○ | ○ |
| The hypothesis will bring no **risks** (2) | ○ | ○ | ○ | ○ | ○ |
| I will **trade my place** with a participant in the hypothesis (3) | ○ | ○ | ○ | ○ | ○ |

---

Q1.6 Can the potential benefits, risks of the hypothesis be evaluated clearly by the above statements?

○ Yes (1)

○ No (2)

---

Q1.7 Are there any additional items that should be included to measure the potential benefits, risks of the hypothesis?

________________________________________________________________

________________________________________________________________

________________________________________________________________

________________________________________________________________

________________________________________________________________

Q1.8 Do you have any additional comments about the evaluation of the potential benefits, risks of the hypothesis?

________________________________________________________________

________________________________________________________________

________________________________________________________________

________________________________________________________________

________________________________________________________________

**End of Block: Default Question Block**

**Start of Block: Block 7**

Example hypothesis:
1) Drug A and drug B are similar in treating disease C. After analyzing data from 2017 in institution X; we notice Drug A was associated with adverse cardiovascular events at a rate significantly higher than with Drug B.

a. Hypothesis: Analysis of date from 2017 in institution Y will show similar higher associations with disease C for Drug A than Drug B.

Q2.1 The significance of this hypothesis will be evaluated by the following metrics (the metrics are for text display purpose, no need to answer):

| | Strongly disagree (1) | Disagree (2) | Neutral (3) | Agree (4) | Strongly agree (5) |
|---|---|---|---|---|---|
| The hypothesis will impact on the **target population** positively (1) | ○ | ○ | ○ | ○ | ○ |
| The hypothesis will reduce the **cost** of the clinical care (e.g.,treatment) (2) | ○ | ○ | ○ | ○ | ○ |
| The hypothesis will impact the **future direction** of clinical care positively (3) | ○ | ○ | ○ | ○ | ○ |

---

Q2.2 Can the significance of the hypothesis be evaluated clearly by the above statements?

○ Yes (1)

○ No (2)

---

Q2.3 Are there any additional items that should be included to measure the significance of the hypothesis?

________________________________________________________________

________________________________________________________________

________________________________________________________________

________________________________________________________________

________________________________________________________________

Q2.4 Do you have any additional comments about the evaluation of the significance of the hypothesis?

Q2.5 The novelty of this hypothesis will be evaluated by the following metrics (the metrics are for text display purpose, no need to answer):

| | Strongly disagree (1) | Disagree (2) | Neutral (3) | Agree (4) | Strongly agree (5) |
| --- | --- | --- | --- | --- | --- |
| The hypothesis can lead to **new findings** (1) | ○ | ○ | ○ | ○ | ○ |
| The hypothesis can alter **previous findings** (2) | ○ | ○ | ○ | ○ | ○ |
| The hypothesis can lead to **novel medical knowledge** (3) | ○ | ○ | ○ | ○ | ○ |
| The hypothesis can lead to **innovation in medical practice** (4) | ○ | ○ | ○ | ○ | ○ |
| The hypothesis can lead to **innovation methodology for research** (5) | ○ | ○ | ○ | ○ | ○ |

Q2.6 Can the novelty of the hypothesis be evaluated clearly by the above statements?

○ Yes (1)

○ No (2)

Q2.7 Are there any additional items that should be included to measure the novelty of the hypothesis?

________________________________________________________________

________________________________________________________________

________________________________________________________________

________________________________________________________________

________________________________________________________________

Q2.8 Do you have any additional comments about the evaluation of the novelty of the hypothesis?

________________________________________________________________

________________________________________________________________

________________________________________________________________

________________________________________________________________

________________________________________________________________

**End of Block: Block 7**

**Start of Block: Block 10**

<u>Example hypothesis</u>:

1) Drug A and drug B are similar in treating disease C. After analyzing data from 2017 in institution X; we notice Drug A was associated with adverse cardiovascular events at a rate significantly higher than with Drug B.

a. Hypothesis: Analysis of date from 2017 in institution Y will show similar higher associations with disease C for Drug A than Drug B.

Q3.1 The clinical relevance of this hypothesis will be evaluated by the following metrics (the metrics are for text display purpose, no need to answer):

| | Strongly disagree (1) | Disagree (2) | Neutral (3) | Agree (4) | Strongly agree (5) |
|---|---|---|---|---|---|
| The hypothesis can impact our understanding of **medical knowledge** (1) | ○ | ○ | ○ | ○ | ○ |
| The hypothesis can impact **clinical practice** (2) | ○ | ○ | ○ | ○ | ○ |
| The hypothesis can impact **health policy** (3) | ○ | ○ | ○ | ○ | ○ |

---

Q3.2 Can the clinical relevance of the hypothesis be evaluated clearly by the above statements?

○ Yes (1)

○ No (2)

---

Q3.3 Are there any additional items that should be included to measure the clinical relevance of the hypothesis?

________________________________________________________________

________________________________________________________________

________________________________________________________________

________________________________________________________________

________________________________________________________________

Q3.4 Do you have any additional comments about the evaluation of the clinical relevance of the hypothesis?

________________________________________________________________

________________________________________________________________

________________________________________________________________

________________________________________________________________

________________________________________________________________

Q3.5 The feasibility of this hypothesis will be evaluated by the following metrics (the metrics are for text display purpose, no need to answer):

| | Strongly disagree (1) | Disagree (2) | Neutral (3) | Agree (4) | Strongly agree (5) |
|---|---|---|---|---|---|
| The hypothesis is feasible to implement regarding **cost** (1) | ○ | ○ | ○ | ○ | ○ |
| The hypothesis is feasible to implement regarding **time** (2) | ○ | ○ | ○ | ○ | ○ |
| The hypothesis is feasible to implement regarding **scope** (3) | ○ | ○ | ○ | ○ | ○ |

Q3.6 How many sub-problems does this hypothesis include? (the metrics are for text display purpose, no need to answer)

________________________________________________________________

Q3.7 Can the feasibility of the hypothesis be evaluated clearly by the above statements?

○ Yes (1)

○ No (2)

Q3.8 Are there any additional items that should be included to measure the feasibility of the hypothesis?

________________________________________________________________

________________________________________________________________

________________________________________________________________

________________________________________________________________

________________________________________________________________

Q3.9 Do you have any additional comments about the evaluation of the feasibility of the hypothesis?

________________________________________________________________

________________________________________________________________

________________________________________________________________

________________________________________________________________

________________________________________________________________

**End of Block: Block 10**

**Start of Block: Block 6**

Example hypothesis:
1) Drug A and drug B are similar in treating disease C. After analyzing data from 2017 in institution X; we notice Drug A was associated with adverse cardiovascular events at a rate significantly higher than with Drug B.

a. Hypothesis: Analysis of date from 2017 in institution Y will show similar higher associations with disease C for Drug A than Drug B.

Q4.1 The testability of this hypothesis will be evaluated by the following metrics (the metrics are for text display purpose, no need to answer):

| | Strongly disagree (1) | Disagree (2) | Neutral (3) | Agree (4) | Strongly agree (5) |
|---|---|---|---|---|---|
| The hypothesis can be **tested** (1) | ○ | ○ | ○ | ○ | ○ |

Q4.2 Can the testability of the hypothesis be evaluated clearly by the above statements?

○ Yes (1)

○ No (2)

Q4.3 Are there any additional items that should be included to measure the testability of the hypothesis?

________________________________________________________________

________________________________________________________________

________________________________________________________________

________________________________________________________________

________________________________________________________________

Q4.4 Do you have any additional comments about the evaluation of the testability of the hypothesis?

________________________________________________________________

________________________________________________________________

________________________________________________________________

________________________________________________________________

________________________________________________________________

Q4.5 The clarity of this hypothesis will be evaluated by the following metrics (the metrics are for text display purpose, no need to answer):

| | Strongly disagree (1) | Disagree (2) | Neutral (3) | Agree (4) | Strongly agree (5) |
|---|---|---|---|---|---|
| The hypothesis provides clear **purpose**(s) (1) | ○ | ○ | ○ | ○ | ○ |
| The hypothesis identifies **focused group(**s) (2) | ○ | ○ | ○ | ○ | ○ |
| The hypothesis specifies **variable**(s) (3) | ○ | ○ | ○ | ○ | ○ |
| The hypothesis specifies **the relationship**(s) between the variables under investigation (4) | ○ | ○ | ○ | ○ | ○ |

Q4.6 Can the clarity of the hypothesis be evaluated clearly by the above statements?

○ Yes (1)

○ No (2)

---

Q4.7 Are there any additional items that should be included to measure the clarity of the hypothesis?

________________________________________________________________

________________________________________________________________

________________________________________________________________

________________________________________________________________

________________________________________________________________

---

Q4.8 Do you have any additional comments about the evaluation of the clarity of the hypothesis?

________________________________________________________________

________________________________________________________________

________________________________________________________________

________________________________________________________________

________________________________________________________________

**End of Block: Block 6**

---

**Start of Block: Block 9**

Example hypothesis:
1) Drug A and drug B are similar in treating disease C. After analyzing data from 2017 in institution X; we notice Drug A was associated with adverse cardiovascular events at a rate significantly higher than with Drug B.

a. Hypothesis: Analysis of date from 2017 in institution Y will show similar higher associations with disease C for Drug A than Drug B.

Q5.1 The ethicality of this hypothesis will be evaluated by the following metrics (the metrics are for text display purpose, no need to answer):

| | Strongly disagree (1) | Disagree (2) | Neutral (3) | Agree (4) | Strongly agree (5) |
|---|---|---|---|---|---|
| There are no **ethical** concerns in this hypothesis (1) | ○ | ○ | ○ | ○ | ○ |

Q5.2 Can the ethicality of the hypothesis be evaluated clearly by the above statements?

○ Yes (1)

○ No (2)

Q5.3 Are there any additional items that should be included to measure the ethicality of the hypothesis?

________________________________________________________________

________________________________________________________________

________________________________________________________________

________________________________________________________________

________________________________________________________________

Q5.4 Do you have any additional comments about the evaluation of the ethicality of the hypothesis?

________________________________________________________________

________________________________________________________________

________________________________________________________________

________________________________________________________________

________________________________________________________________

---

Q5.5 The interesting of this hypothesis will be evaluated by the following metrics (the metrics are for text display purpose, no need to answer):

| | Strongly disagree (1) | Disagree (2) | Neutral (3) | Agree (4) | Strongly agree (5) |
|---|---|---|---|---|---|
| This hypothesis **interests** me (1) | ○ | ○ | ○ | ○ | ○ |
| I will **pursue** the hypothesis if possible (2) | ○ | ○ | ○ | ○ | ○ |

---

Q5.6 Can the interest of the hypothesis be evaluated clearly by the above statements?

○ Yes (1)

○ No (2)

---

Q5.7 Are there any additional items that should be included to measure the interesting of the hypothesis?

________________________________________________________________

________________________________________________________________

Q5.8 Do you have any additional comments about the evaluation of the interesting of the hypothesis?

________________________________________________________________

________________________________________________________________

________________________________________________________________

________________________________________________________________

________________________________________________________________

**End of Block: Block 9**

---

**Start of Block: Block 8**

Example hypothesis:
1) Drug A and drug B are similar in treating disease C. After analyzing data from 2017 in institution X; we notice Drug A was associated with adverse cardiovascular events at a rate significantly higher than with Drug B.

a. Hypothesis: Analysis of date from 2017 in institution Y will show similar higher associations with disease C for Drug A than Drug B.

Q6.1 The overall quality score of the hypothesis on each dimension (1--the lowest; 5--the highest) will be evaluated by the following metrics (the metrics are for text display purpose, no need to answer):

| | | | | | | | | | |
|---|---|---|---|---|---|---|---|---|---|
| 1 | 1 | 2 | 2 | 3 | 3 | 3 | 4 | 4 | 5 |

| |
|---|
| Validity () |
| Significance () |
| Novelty () |
| Clinical relevance () |
| Feasibility () |
| Testability () |
| Clarity () |
| Ethicality () |
| Potential benefits and risks () |
| Interesting () |

Q6.2 Do you have any additional comments about the overall quality score evaluation of the hypothesis?

________________________________________________________________

________________________________________________________________

________________________________________________________________

________________________________________________________________

________________________________________________________________

Q6.3 Additional comments on this hypothesis metrics:

________________________________________________________________

________________________________________________________________

________________________________________________________________

End of Block: Block 8
