## Supplementary material for "Development, validation, and usage of metrics to evaluate the quality of clinical research hypotheses": A survey (Appendix 2) that served as the medium validation instrument was used among all expert panel members

### Metrics_Expert_Feedback_20221005

Start of Block: Default Question Block

This is the third round of external feedback surveys about the metrics (i.e., instrument) you will use to evaluate each hypothesis generated by VIADS experiment participants. This time, we added one more option as an overall assessment under each dimension instead of using the subitems. Please tell us what you think, an overall assessment or more detailed subitems.

Example hypothesis to provide some basic idea about a hypothesis. Our instrument is intended to be used to evaluate all hypotheses within a clinical research context:
1) Drug A and drug B are similar in treating disease C. After analyzing data from 2017 in institution X, we noticed Drug A was associated with adverse cardiovascular events at a rate significantly higher than with Drug B.

a. Hypothesis: Data analysis from 2017 in institution Y will show similar higher associations with disease C for Drug A than Drug B.

Q1.1 The following metrics are intended to evaluate the **validity** of any given hypothesis. This is an overall judgment based on your knowledge and experience. Please select the "Unnecessary subitem" or "Use this one only".

| | Strongly disagree (1) | Disagree (2) | Neutral (3) | Agree (4) | Strongly agree (5) | Unable to assess (6) | Unnecessary subitem (7) | Use this one only (8) |
|---|---|---|---|---|---|---|---|---|
| The hypothesis is valid **scientifically** (1) | ○ | ○ | ○ | ○ | ○ | ○ | ○ | ○ |
| The hypothesis is valid **clinically**, i.e., sound basis clinically (2) | ○ | ○ | ○ | ○ | ○ | ○ | ○ | ○ |
| The hypothesis is **valid** (i.e., use this as an overall one instead of the above two subitems) (3) | ○ | ○ | ○ | ○ | ○ | ○ | ○ | ○ |

---

Q1.2 Can the validity of the hypothesis being evaluated by the above metrics?

○ Yes (1)

○ No (2)

---

Q1.3 Are there any additional items that should be included to measure the validity of a given hypothesis?

________________________________________________________________

Q1.4 Do you have any additional comments about evaluating the validity of a given hypothesis?

Q1.5 The potential benefits and risks for stakeholders, who will be the beneficiaries if this hypothesis can be translated into a large-scale study, will be evaluated by the following metrics. Please select the "Unnecessary subitem" or "Use this one only".

| | Strongly disagree (1) | Disagree (2) | Neutral (3) | Agree (4) | Strongly agree (5) | Unable to assess (6) | Unnecessary subitem (7) | Use this one only (8) |
|---|---|---|---|---|---|---|---|---|
| The successful testing of this hypothesis will bring significant **benefits** to targeted audiences (e.g., patients, providers) (1) | ○ | ○ | ○ | ○ | ○ | ○ | ○ | ○ |
| The testing of this hypothesis will bring no **risks** or tolerable risks to targeted audiences considering the benefits (2) | ○ | ○ | ○ | ○ | ○ | ○ | ○ | ○ |
| The successful testing of this hypothesis will bring significant **benefits** to targeted audiences (e.g., patients, providers), i.e., the | ○ | ○ | ○ | ○ | ○ | ○ | ○ | ○ |

| |
|---|
| overall benefits outweigh the **risks** (4) |

Q1.6 Can the potential benefits and risks of the stakeholders who participate in testing this hypothesis be evaluated by the above statements?

- Yes (1)
- No (2)

Q1.7 Are there any additional items that should be included to measure the potential benefits and risks of the stakeholders when testing this hypothesis?

________________________________________________________________

________________________________________________________________

________________________________________________________________

________________________________________________________________

________________________________________________________________

Q1.8 Do you have any additional comments about the evaluation of the potential benefits and risks of potential stakeholders while testing this hypothesis?

________________________________________________________________

________________________________________________________________

________________________________________________________________

________________________________________________________________

________________________________________________________________

End of Block: Default Question Block

Start of Block: Block 7

Example hypothesis:
1) Drug A and drug B are similar in treating disease C. After analyzing data from 2017 in institution X, we noticed Drug A was associated with adverse cardiovascular events at a rate significantly higher than with Drug B.

a. Hypothesis: Data analysis from 2017 in institution Y will show similar higher associations with disease C for Drug A than Drug B.

Q2.1 The following metrics will evaluate the **significance** of this hypothesis, i.e., the significance of a study used to test this hypothesis. Please select the "Unnecessary subitem" or "Use this one only".

| | Strongly disagree (1) | Disagree (2) | Neutral (3) | Agree (4) | Strongly agree (5) | Unable to assess (6) | Unnecessary subitem (7) | Use this one only (8) |
|---|---|---|---|---|---|---|---|---|
| The hypothesis focuses on addressing **established medical needs**, e.g., a major medical problem affecting a relatively large population or the potential magnitude of improvement via testing this hypothesis for a severe condition (4) | ○ | ○ | ○ | ○ | ○ | ○ | ○ | ○ |
| The test results of this hypothesis have the potential to impact the **future direction** of clinical practice positively (3) | ○ | ○ | ○ | ○ | ○ | ○ | ○ | ○ |
| The test results of this hypothesis have the | ○ | ○ | ○ | ○ | ○ | ○ | ○ | ○ |

| | | | | | | | | |
|---|---|---|---|---|---|---|---|---|
| potential to impact the **target population** positively on average (1) | | | | | | | | |
| The test of this hypothesis will be a worthwhile effort regarding the **cost** and benefit (2) | ○ | ○ | ○ | ○ | ○ | ○ | ○ | ○ |
| Overall, this hypothesis is **significant** considering medical needs, cost and benefits, target population, and future directions of clinical practice. (7) | ○ | ○ | ○ | ○ | ○ | ○ | ○ | ○ |

---

Q2.2 Can the significance of a study that tests any given hypothesis be evaluated by the above statements?

○ Yes (1)

○ No (2)

---

Q2.3 Are there any additional items that should be included to measure the significance of a study that tests any given hypothesis?

________________________________________________________________

Q2.4 Do you have any additional comments about the evaluation of the significance of a study to test any given hypothesis?

Q2.5 The following metrics will evaluate the **novelty** of a study to test any given hypothesis. Please select the "Unnecessary subitem" or "Use this one only".

| | Strongly disagree (1) | Disagree (2) | Neutral (3) | Agree (4) | Strongly agree (5) | Unable to assess (6) | Unnecessary subitem (7) | Use this one only (8) |
|---|---|---|---|---|---|---|---|---|
| The test of a given hypothesis can lead to **innovation in medical practice** (4) | ○ | ○ | ○ | ○ | ○ | ○ | ○ | ○ |
| The test of a given hypothesis can lead to **innovation methodology for clinical research** (5) | ○ | ○ | ○ | ○ | ○ | ○ | ○ | ○ |
| The test of a given hypothesis can alter **previous findings**, i.e., has the potential to bring in paradigm shift in the field (2) | ○ | ○ | ○ | ○ | ○ | ○ | ○ | ○ |
| The test of a given hypothesis can lead to **novel medical knowledge** (3) | ○ | ○ | ○ | ○ | ○ | ○ | ○ | ○ |
| The test of a given hypothesis can lead to **new findings**, | ○ | ○ | ○ | ○ | ○ | ○ | ○ | ○ |

| | | | | | | | | |
|---|---|---|---|---|---|---|---|---|
| which can be incremental (1) | | | | | | | | |
| Overall, this hypothesis is **novel** (6) | ○ | ○ | ○ | ○ | ○ | ○ | ○ | ○ |

Q2.6 Can the novelty of a study to test a given hypothesis be evaluated by the above statements?

○ Yes (1)

○ No (2)

Q2.7 Are there any additional items that should be included to measure the novelty of a study to test a given hypothesis?

________________________________________________________________

________________________________________________________________

________________________________________________________________

________________________________________________________________

________________________________________________________________

Q2.8 Do you have any additional comments about the evaluation of the novelty of a study to test a given hypothesis?

________________________________________________________________

________________________________________________________________

________________________________________________________________

________________________________________________________________

________________________________________________________________

End of Block: Block 7

Start of Block: Block 10

<u>Example hypothesis</u>:

1) Drug A and drug B are similar in treating disease C. After analyzing data from 2017 in institution X, we noticed Drug A was associated with adverse cardiovascular events at a rate significantly higher than with Drug B.

a. Hypothesis: Data analysis from 2017 in institution Y will show similar higher associations with disease C for Drug A than Drug B.

Q3.1 The clinical relevance of a study to test a given hypothesis will be evaluated by the following metrics. Please select the "Unnecessary subitem" or "Use this one only".

| | Strongly disagree (1) | Disagree (2) | Neutral (3) | Agree (4) | Strongly agree (5) | Unable to assess (6) | Unnecessary subitem (7) | Use this one only (8) |
|---|---|---|---|---|---|---|---|---|
| The test of a given hypothesis has the potential to impact **current clinical practice**, including patient safety, care quality (2) | ○ | ○ | ○ | ○ | ○ | ○ | ○ | ○ |
| The test of a given hypothesis has the potential to impact our understanding of **medical knowledge** (1) | ○ | ○ | ○ | ○ | ○ | ○ | ○ | ○ |
| The test of a given hypothesis has the potential to impact **health policy** (3) | ○ | ○ | ○ | ○ | ○ | ○ | ○ | ○ |
| Overall, this hypothesis is **clinically relevant** (4) | ○ | ○ | ○ | ○ | ○ | ○ | ○ | ○ |

Q3.2 Can the clinical relevance of a study to test a given hypothesis be evaluated by the above statements?

- ◯ Yes (1)
- ◯ No (2)

---

Q3.3 Are there any additional items that should be included to measure the clinical relevance of a study to test a given hypothesis?

________________________________________________________________

________________________________________________________________

________________________________________________________________

________________________________________________________________

________________________________________________________________

---

Q3.4 Do you have any additional comments about the evaluation of the clinical relevance of a study to test a given hypothesis?

________________________________________________________________

________________________________________________________________

________________________________________________________________

________________________________________________________________

________________________________________________________________

---

Q3.5 The feasibility of conducting a study to test a given hypothesis will be evaluated by the following metrics assuming the budget limit is 5 k US dollars and 0.5 years of a graduate student's time. Please select the "Unnecessary subitem" or "Use this one only".

| | Strongly disagree (1) | Disagree (2) | Neutral (3) | Agree (4) | Strongly agree (5) | Unable to assess (6) | Unnecessary subitem (7) | Use this one only (8) |
|---|---|---|---|---|---|---|---|---|
| A study to test a given hypothesis is feasible regarding **needed cost**, i.e., needed resources or tools (1) | ○ | ○ | ○ | ○ | ○ | ○ | ○ | ○ |
| A study to test a given hypothesis is feasible regarding **needed time** to conduct the study and to follow up (2) | ○ | ○ | ○ | ○ | ○ | ○ | ○ | ○ |
| A study to test a given hypothesis is feasible regarding **scope**, i.e., a well-defined question (3) | ○ | ○ | ○ | ○ | ○ | ○ | ○ | ○ |
| Overall, this hypothesis is **feasible** | ○ | ○ | ○ | ○ | ○ | ○ | ○ | ○ |

to test (4)

Q3.6 How many sub-problems does this hypothesis include, if there are any?

________________________________________________________________

Q3.7 Can the feasibility of a study to test a given hypothesis be evaluated by the above statements?

- ○ Yes (1)
- ○ No (2)

Q3.8 Are there any additional items that should be included to measure the feasibility of a study to test a given hypothesis?

________________________________________________________________

________________________________________________________________

________________________________________________________________

________________________________________________________________

________________________________________________________________

Q3.9 Do you have any additional comments about the evaluation of the feasibility of a study to test a given hypothesis?

________________________________________________________________

________________________________________________________________

________________________________________________________________

________________________________________________________________

End of Block: Block 10

Start of Block: Block 6

Example hypothesis:
1) Drug A and drug B are similar in treating disease C. After analyzing data from 2017 in institution X, we noticed Drug A was associated with adverse cardiovascular events at a rate significantly higher than with Drug B.

a. Hypothesis: Data analysis from 2017 in institution Y will show similar higher associations with disease C for Drug A than Drug B.

Q4.1 The testability of a given hypothesis will be evaluated by the following metrics. Please select the "Unnecessary subitem" or "Use this one only".

| | Strongly disagree (1) | Disagree (2) | Neutral (3) | Agree (4) | Strongly agree (5) | Unable to assess (6) | Unnecessary subitem (7) | Use this one only (8) |
|---|---|---|---|---|---|---|---|---|
| The hypothesis can be **tested** in an ideal setting (1) | ○ | ○ | ○ | ○ | ○ | ○ | ○ | ○ |
| There are an adequate **number of patients** to choose from to participate in a study to test a given hypothesis (2) | ○ | ○ | ○ | ○ | ○ | ○ | ○ | ○ |
| Overall, this hypothesis is **testable** (3) | ○ | ○ | ○ | ○ | ○ | ○ | ○ | ○ |

Q57 How many assumptions does this hypothesis include, if there are any?

________________________________________________________________

Q58 Are some or all these assumptions stringent?

- ◯ Yes (1)
- ◯ No (2)
- ◯ Not sure (3)

---

Q4.2 Can the testability of a given hypothesis be evaluated by the above statements?

- ◯ Yes (1)
- ◯ No (2)

---

Q4.3 Are there any additional items that should be included to measure the testability of any given hypothesis?

________________________________________________________________

________________________________________________________________

________________________________________________________________

________________________________________________________________

________________________________________________________________

---

Q4.4 Do you have any additional comments about evaluating the testability of any given hypothesis?

________________________________________________________________

________________________________________________________________

________________________________________________________________

________________________________________________________________

________________________________________________________________

Q4.5 The clarity of a given hypothesis will be evaluated by the following metrics. Please select the "Unnecessary subitem" or "Use this one only".

| | Strongly disagree (1) | Disagree (2) | Neutral (3) | Agree (4) | Strongly agree (5) | Unable to assess (6) | Unnecessary subitem (7) | Use this one only (8) |
|---|---|---|---|---|---|---|---|---|
| The hypothesis provides clear **purpose**(s) (1) | ○ | ○ | ○ | ○ | ○ | ○ | ○ | ○ |
| The hypothesis identifies **focused group(**s) (2) | ○ | ○ | ○ | ○ | ○ | ○ | ○ | ○ |
| The hypothesis specifies **variable**(s) (3) | ○ | ○ | ○ | ○ | ○ | ○ | ○ | ○ |
| The hypothesis specifies **the relationship**(s) between the variables under investigation (4) | ○ | ○ | ○ | ○ | ○ | ○ | ○ | ○ |
| Overall, this hypothesis is **clear** (5) | ○ | ○ | ○ | ○ | ○ | ○ | ○ | ○ |

Q4.6 Can the clarity of a given hypothesis be evaluated by the above statements?

○ Yes (1)

○ No (2)

Q4.7 Are there any additional items that should be included to measure the clarity of a given hypothesis?

________________________________________________________________

________________________________________________________________

________________________________________________________________

________________________________________________________________

________________________________________________________________

Q4.8 Do you have any additional comments about evaluating the clarity of a given hypothesis?

________________________________________________________________

________________________________________________________________

________________________________________________________________

________________________________________________________________

________________________________________________________________

**End of Block: Block 6**

**Start of Block: Block 9**

Example hypothesis:
1) Drug A and drug B are similar in treating disease C. After analyzing data from 2017 in institution X, we noticed Drug A was associated with adverse cardiovascular events at a rate significantly higher than with Drug B.

a. Hypothesis: Data analysis from 2017 in institution Y will show similar higher associations with disease C for Drug A than Drug B.

Q5.1 The following metrics will evaluate the **ethicality** of a study to test a given hypothesis. Please select the "Unnecessary subitem" or "Use this one only".

| | Yes (1) | No (2) | Unable to assess (3) | Unnecessary subitem (6) | Use this one only (7) |
|---|---|---|---|---|---|
| ⊗ There are no **ethical** concerns when conducting a study to test a given hypothesis, i.e., regarding patients, investigators, providers, and the conduction of the study (1) | ○ | ○ | ○ | ○ | ○ |
| ⊗ I will **trade my place** with a participant without hesitation in a study to test a given hypothesis (2) | ○ | ○ | ○ | ○ | ○ |
| Overall, it is **ethical** to test this hypothesis (3) | ○ | ○ | ○ | ○ | ○ |

Q5.2 Can the ethicality of a study to test a given hypothesis be evaluated by the above statements?

- Yes (1)
- No (2)

---

Q5.3 Are there any additional items that should be included to measure the ethicality of a study to test a given hypothesis?

________________________________________________________________

________________________________________________________________

________________________________________________________________

________________________________________________________________

________________________________________________________________

---

Q5.4 Do you have any additional comments about the evaluation of the ethicality of a study to test a given hypothesis?

________________________________________________________________

________________________________________________________________

________________________________________________________________

________________________________________________________________

________________________________________________________________

---

Q5.5 The interestingness of a given hypothesis will be evaluated by the following metrics. Please select the "Unnecessary subitem" or "Use this one only".

| | Strongly disagree (1) | Disagree (2) | Neutral (3) | Agree (4) | Strongly agree (5) | Unable to assess (6) | Unnecessary subitem (7) | Use this one only (8) |
|---|---|---|---|---|---|---|---|---|
| This hypothesis **interests** me (1) | ○ | ○ | ○ | ○ | ○ | ○ | ○ | ○ |
| I will **pursue** the hypothesis if possible/feasible (2) | ○ | ○ | ○ | ○ | ○ | ○ | ○ | ○ |
| Overall, this is an **interesting** (i.e., the researcher should be able to find collaborators easily) hypothesis (3) | ○ | ○ | ○ | ○ | ○ | ○ | ○ | ○ |

Q5.6 Can the interestingness of a study to test a given hypothesis be evaluated by the above statements?

○ Yes (1)

○ No (2)

Q5.7 Are there any additional items that should be included to measure the interestingness of a study to test a given hypothesis?

________________________________________________________________

________________________________________________________________

________________________________________________________________

________________________________________________________________

Q5.8 Do you have any additional comments about the evaluation of the interestingness of the study to test a given hypothesis?

________________________________________________________________

________________________________________________________________

________________________________________________________________

________________________________________________________________

________________________________________________________________

**End of Block: Block 9**

---

**Start of Block: Block 8**

Example hypothesis:
1) Drug A and drug B are similar in treating disease C. After analyzing data from 2017 in institution X, we noticed Drug A was associated with adverse cardiovascular events at a rate significantly higher than with Drug B.

a. Hypothesis: Data analysis from 2017 in institution Y will show similar higher associations with disease C for Drug A than Drug B.

Q6.1 The overall quality score of the hypothesis on each dimension (1--the lowest; 5--the highest) will be evaluated by the following metrics:

Not Applicable

1 1 2 2 3 3 3 4 4 5

| |
|---|
| Validity () |
| Significance () |
| Novelty () |
| Clinical relevance () |
| Feasibility () |
| Testability () |
| Clarity () |
| Ethicality () |
| Potential benefits and risks () |
| Interesting () |

Q6.2 Do you have any additional comments about the overall quality score evaluation of a given hypothesis?

Q6.3 Additional comments on this hypothesis metrics:

End of Block: Block 8
