## Supplementary material for "Development, validation, and usage of metrics to evaluate the quality of clinical research hypotheses": We present comprehensive (10 items and 39 subitems, Appendix 3) and brief versions (3 items, 12 subitems, Table 1, Appendix 4)

### 3-item clinical hypotheses evaluation

Overall guidance for the survey:

- If you hover the mouse over each dimension, such as Validity, you will see the definition and examples of the dimension.
- Example hypothesis 1 with an overall 4.0 rating: Given the COVID-19 pandemic, more patients use telemedicine for their healthcare services. We hypothesize that after COVID-19, more patients would like to have telehealth visits
- Example hypothesis 2 with a 4.0 rating: Patients who have hypertension between 2005 and 2015, do hypertension patients have a higher rate of morbid obesity (ICD9 codes: 27801) in 2015 than in 2005?
- Example hypothesis 3 with an overall 2.8 rating: Some kind of respiratory disorder may lead to the COPD
- Example hypothesis 4 with 2.47 rating: I hypothesize that the different infant mortality rates among private health insurance beneficiaries and Medicaid beneficiaries will help us infer the social-economic status differences between the two groups to identify the key factors contributing to the higher infant mortality rate.
- **To combine 2005 and 2015 data sets and treat the combined data set as a whole and look at it more closely** is not a hypothesis; if a statement is ***not a hypothesis***, please select the lowest score for validity and ***no other dimensions need to be evaluated for that hypothesis***.

Hypothesis 1: To compare different states in the USA if the incidence of 5859 (ICD9 code: chronic kidney diseases, unspecified) is correlated to kidney transplantation surgeries. If there are differences, what causes these differences? Donors? Surgeons? Or other reasons.

Please provide an overall score from 1 (the lowest) to 5 (the highest) on each dimension for the hypothesis.

| | 1 | 2 | 3 | 4 | 5 | Not Applicable |
|---|---|---|---|---|---|---|
| Validity | | | | | | ☐ |
| Significance | | | | | | ☐ |
| Feasibility | | | | | | ☐ |

Powered by Qualtrics
